# Diagnosis coding of Chronic Kidney Disease in Type 2 Diabetes in UK primary care

**DOI:** 10.1101/2023.05.11.23289836

**Authors:** Rose Sisk, Rory Cameron, Waqas Tahir, Camilla Sammut-Powell

## Abstract

1.

**Background:** Type 2 diabetes (T2D) is a leading cause of both chronic kidney disease (CKD) and onward progression to end stage renal disease. Timely diagnosis coding of CKD in patients with T2D could lead to improvements in quality of care and patient outcomes.

**Aim:** To assess the consistency between estimated glomerular filtration rate (eGFR) based evidence of CKD and CKD diagnosis coding in UK primary care.

**Design and Setting:** A retrospective analysis of electronic health record data in a cohort of people with type 2 diabetes from 60 primary care centres within England between 2012 and 2022.

**Method:** We estimated the incidence rate of CKD per 100 person-years using eGFR-based CKD and diagnosis codes. Logistic regression was applied to establish which attributes were associated with diagnosis coding. Time from eGFR-based CKD to entry of a diagnosis code was summarised using the median and interquartile range.

**Results:** The overall incidence of CKD was 2.32 (95% CI: 2.24, 2.41) and significantly different between eGFR-based criteria and diagnosis codes: 1.98 (95% CI: 1.90, 2.05) vs 1.06 (95% CI: 1.00, 1.11) respectively; p<0.001. Only 46% of CKD incidences identified using eGFR-based criteria had a corresponding diagnosis code. Younger patients, patients with a higher severity CKD stage, and patients with an observed urine-albumin-to-creatinine ratio were more likely to have a diagnosis code.

**Conclusion:** Diagnosis coding of patients with eGFR-based evidence of CKD in UK primary care is poor within patients with type 2 diabetes, despite CKD being a well-known complication of diabetes.

**How this fits in:** Type 2 diabetes is a recognised cause of chronic kidney disease (CKD), and early identification and management of CKD can reduce the risk of progression and related complications. Diagnosis coding of CKD is associated with better patient outcomes, yet we have observed that less than half of patients with type 2 diabetes who meet eGFR-based criteria for stage 3-5 CKD have a CKD diagnosis code in their primary care record. There is a need to understand why CKD diagnosis coding practices are subpar in primary care and this research acts as a call-to-action to improve.

## 3. Background

Chronic kidney disease (CKD) is a chronic, progressive condition that places an enormous burden on healthcare systems and patients globally (1). Type 2 diabetes (T2D) is recognised as a leading cause of CKD and is associated with rapid deterioration in kidney function and an increased risk of end stage renal disease (ESRD)(2,3), and both CKD and T2D are risk factors for cardiovascular complications (4). It is therefore essential that people with type 2 diabetes are regularly screened using key markers of kidney health to facilitate early detection and appropriate management of CKD(5,6).

CKD is characterised by a sustained drop in glomerular filtration rate, often evidenced by a reduced estimated glomerular filtration rate (eGFR), and/or persistent albuminuria(7). The combination of eGFR, calculated using serum creatinine, and albuminuria, with severity characterised using the urine albumin-to-creatinine ratio (UACR), are used to classify the disease and provide a patient’s estimated risk of progression(7). As such, existing guidelines from the American Diabetes Association (ADA)(8) and European Association of the Study of Diabetes (EASD)(9) dictate that people with T2D should receive serum creatinine and urinary albumin tests annually. In the UK, the National Institute for Health and Care Excellence promotes the implementation of the “nine key care processes” for the management of T2D. These care processes form the foundation of annual reviews that are undertaken within primary care, and include the measurement of both serum creatinine and urinary albumin. However, albumin testing uptake remains extremely poor compared to serum creatinine testing (10).

Early identification of CKD within the primary care setting can facilitate earlier intervention to slow down the progression of kidney disease and reduce the risk of further complications. Such interventions may include lifestyle advice and targeted risk factor management via pharmaceutical intervention(11,12), including sodium glucose cotransporter-2 inhibitors (SGLT2is) that have recently shown promise in improving renal and cardiovascular outcomes in patients with and without T2D(13–15).

Although CKD is generally well-defined using lab-derived measures (such as eGFR and UACR), the rate of diagnosis coding of CKD remains poor within UK primary care (16,17), despite the existence of financial incentives that promote identification and management of CKD under the Quality and Outcomes Framework (QOF). Yet, diagnostic coding of CKD in primary care is associated with lower rates of hospitalisation for cardiovascular events (18), and improved quality of care(17). Existing work exploring the rate of diagnosis coding for CKD in UK primary care has focused on the general population, with diabetes considered as a subgroup (16,17). However, people with T2D engage with primary care services more frequently and for different reasons than the general population. It is therefore important to quantify how well CKD is coded specifically in a T2D population, within the context of the NICE, ADA/EASD and QOF guidelines relevant to this group.

This study is a retrospective analysis of routinely collected UK primary care data to establish: the incidence of CKD estimated using eGFR-based evidence and/or diagnosis codes, the proportion of eGFR-based CKD incidences that have a diagnosis code, and the timeliness of CKD diagnosis coding in a cohort of patients with T2D.

## 4. Methods

This is a retrospective cohort study using routinely collected UK primary care data from 60 general practitioner (GP) practices across England between 2012 and 2022.

### 4.1. Cohort

All patients with T2D were considered for inclusion in the study cohort. A burn-in period was defined from the start of data collection (February 2012) to 6th April 2015 to exclude patients with pre-existing CKD (either eGFR-based or coded) prior to the beginning of the study period; any patients with evidence of CKD in their record within the burn-in period or prior to their diagnosis of type 2 diabetes were excluded. The study period ran from the 2015/16 to 2020/21 fiscal years.

### 4.2. Definitions

The eGFR-based cases of CKD were ascertained using repeated serum creatinine or eGFR measurements, with eGFR calculated from serum creatinine using the 2021 CKD-epi formula(19) without racial adjustment. If both an eGFR and serum creatinine measurement existed on the same day for a patient, the serum creatinine measurement was retained and used to calculate eGFR. Patients were classed as having eGFR-based CKD if they had 2 or more eGFR measurements below 60/*ml*/*min*/1.73*m*^2^at least 90 days apart, upto a maximum of 15 months apart. Any eGFR measurements occurring between the dates of those observations were required to have a median of below 60/*ml*/*min*/1.73*m*^2^.

An upper limit on the time between qualifying eGFR measurements was imposed to distinguish between a sustained drop in kidney function and two acute episodes of kidney injury or infection. This upper limit was set to 15 months between measures to allow identification of eGFR-based CKD using measurements obtained at two consecutive diabetic annual reviews.

Diagnosis codes were identified using READ v2, READ CTV3 and SNOMED terminologies. Only CKD diagnoses of stages G3-5 (eGFR < 60) were classed as CKD within this analysis. Codelists for each terminology are provided in the supplementary materials (Tables S**7**, S**8** and S**9**).

### 4.3. Statistical analysis

The number of patients with a diagnosis code and/or meeting the eGFR-based criteria for CKD were estimated for each included fiscal year, to align with the financial incentives and audits within the National Health Service. We further present the number (and percentage %) of patients that have at least one observation of eGFR (or serum creatinine) during that year, and how many of these have at least one eGFR below 60/*ml*/*min*/1.73*m*^2^.

To estimate the annual incidence of CKD, three “at-risk” cohorts of patients were established (one for each definition of CKD) for each fiscal year from 2015/16 to 2020/21. Patients are considered at-risk if they are registered with a participating GP practice with a diagnosis of type 2 diabetes before the end of the fiscal year, and no prior evidence of CKD. Evidence of CKD is defined as having:

1. eGFR-based CKD
2. A CKD diagnosis code
3. eGFR-based CKD or a CKD diagnosis code (composite criteria).

Incidence rates are presented per 100 person-years. Follow-up began at the latest of the beginning of the fiscal year, or their date of type 2 diabetes diagnosis. Follow-up ended at the earliest of death, CKD incidence, deregistration from their GP practice, or the end of the fiscal year. Poisson-based 95% confidence intervals for incidence rates were estimated.

For eGFR-based CKD, we estimated the median time interval between the two qualifying eGFR measurements, and we identified the number and proportion of patients with an observed UACR (and its severity category) in the year prior to their first qualifying eGFR. We quantified the number and proportion of patients with eGFR-based CKD that also received a CKD diagnosis code, and summarised the attributes (age, gender, deprivation, duration of diabetes, eGFR-stage, time between qualifying eGFRs, UACR, and indicators of medication prescribing) of patients with and without CKD diagnosis codes descriptively using the median and interquartile range for continuous measures, and count and percentage for categorical measures. Logistic regression was used to identify which patient attributes were associated with entry of a CKD diagnosis code. Gender, eGFR, UACR, deprivation decile and medication indicators were included as categorical predictors, and age, duration of diabetes, and time between qualifying eGFRs were included as continuous predictors. Deprivation decile is assigned using the postcode of a patient’s GP practice, from 1 = most deprived to 10 = least deprived.

For patients that met the eGFR-based criteria and had a CKD diagnosis code during the follow-up period, we quantified the number and proportion of patients that received a diagnosis code before and after the second qualifying eGFR observation. When a patient met the eGFR-based criteria and had a CKD diagnosis code after their second qualifying measurement, we defined this as clinician-verified CKD. The proportion of patients with eGFR-based CKD that had clinician-verified CKD was estimated and the time to verification was summarised using the median and interquartile range.

All analyses were conducted in R version 4.2.3.

## 5. Results

A total of 32,276 patients were found to have type 2 diabetes and were at risk of CKD (Figure **1**). Of these, 13,945 patients (43%) were receiving care from a GP practice in a highly deprived area (IMD = 1 or 2) and the majority of patients had been first diagnosed with type 2 diabetes in the 2 years prior to study entry (median 1.8 years; IQR 0 – 7 years), with an average follow-up of 5 years (IQR: (2.31, 6.00)) (Table **1**).

**Table 1:** Characteristics of patient cohort at the time of cohort entry

| Variable | Patients with T2D and no previous evidence of CKD stages G3-5 (N = 32,276) |
| --- | --- |
| **Age<sup>1</sup>** | 60 (50, 70) |
| **Gender** |  |
| Female | 13,865 (43%) |
| Male | 18,411 (57%) |
| **Duration of follow-up (years)<sup>1</sup>** | 5.01 (2.31, 6.00) |
| **Duration of diabetes (years)<sup>1</sup>** | 1.8 (0.0, 7.0) |
| **CKD Diagnosis Code** | 1,463 (4.5%) |
| **Time to Diagnosis Code (years)<sup>1</sup>** | 3.10 (1.49, 5.41) |
| **eGFR-based CKD** | 2,667 (8.3%) |
| **Time to Lab-based CKD (years)<sup>1</sup>** | 2.66 (1.26, 4.41) |
| **Lab-based or Coded CKD** | 3,102 (9.6%) |
| **Time to Lab-based or Coded CKD (years)<sup>1</sup>** | 2.52 (1.12, 4.36) |
| **Deprivation decile** |  |
| 1-2 (highly deprived) | 13,945 (43%) |
| 3-4 | 6,070 (19%) |
| 5-6 | 3,309 (10%) |
| 7-10 (least deprived) | 8,952 (28%) |
| **General Practices (N)** | 60 |
| **Number of patients per practice<sup>1</sup>** | 472 (277, 603) |

<sup>1</sup>Median (LQ, UQ)

**Figure 1:**
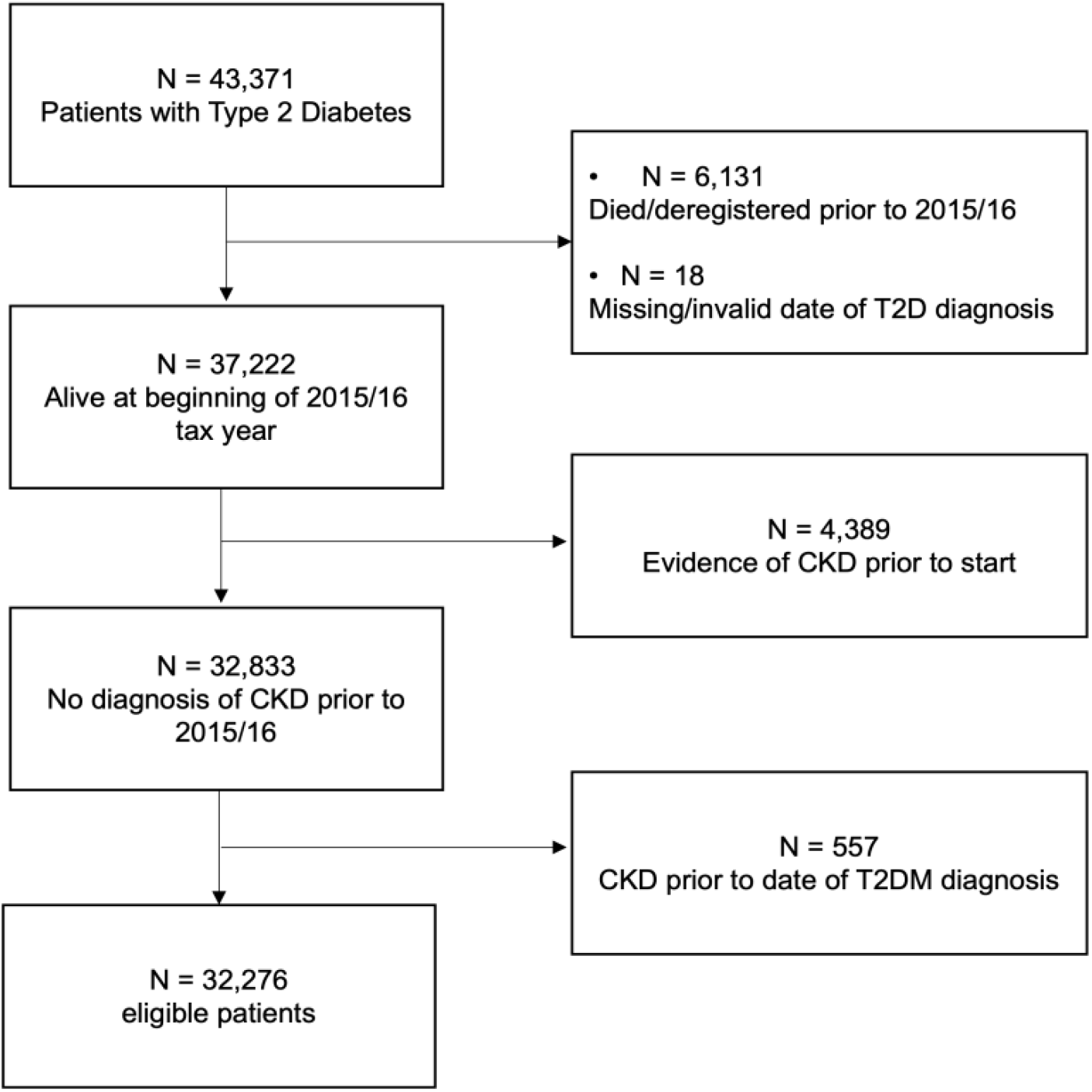
Flowchart of patient inclusions and exclusions

At least one measurement of an eGFR less than 60/*ml*/*min*/1.73*m*^2^was observed in 4,351 patients. However, only 3,000 (68.9%) had a follow-up eGFR (serum creatinine) within 6 months of their first abnormal result (supplementary figure S3).

### 5.1. CKD incidence

Over the study period, the incidence rate of CKD was 2.32 (95% CI: 2.24, 2.41) per 100 person-years of follow-up, using the composite definition of eGFR-based criteria or presence of a CKD diagnosis code. The CKD incidence estimates using eGFR-based criteria and diagnosis codes separately were 1.98 (95% CI: 1.90, 2.05) and 1.06 (95% CI: 1.00, 1.11) respectively. The combined incidence estimate across all fiscal years was significantly higher in the composite criteria than either the eGFR-based criteria (p<0.001) or diagnosis code criteria (p<0.001). Incidence estimates using only CKD diagnosis codes significantly underestimated the overall incidence, as estimated using the composite criteria, across all annual estimates. Incidence estimates using only the eGFR-based criteria were not significantly different from the composite criteria in fiscal years 2018/19, 2019/20 and 2020/21 (Figure 2A, supplementary Table S**5**).

**Figure 2:**
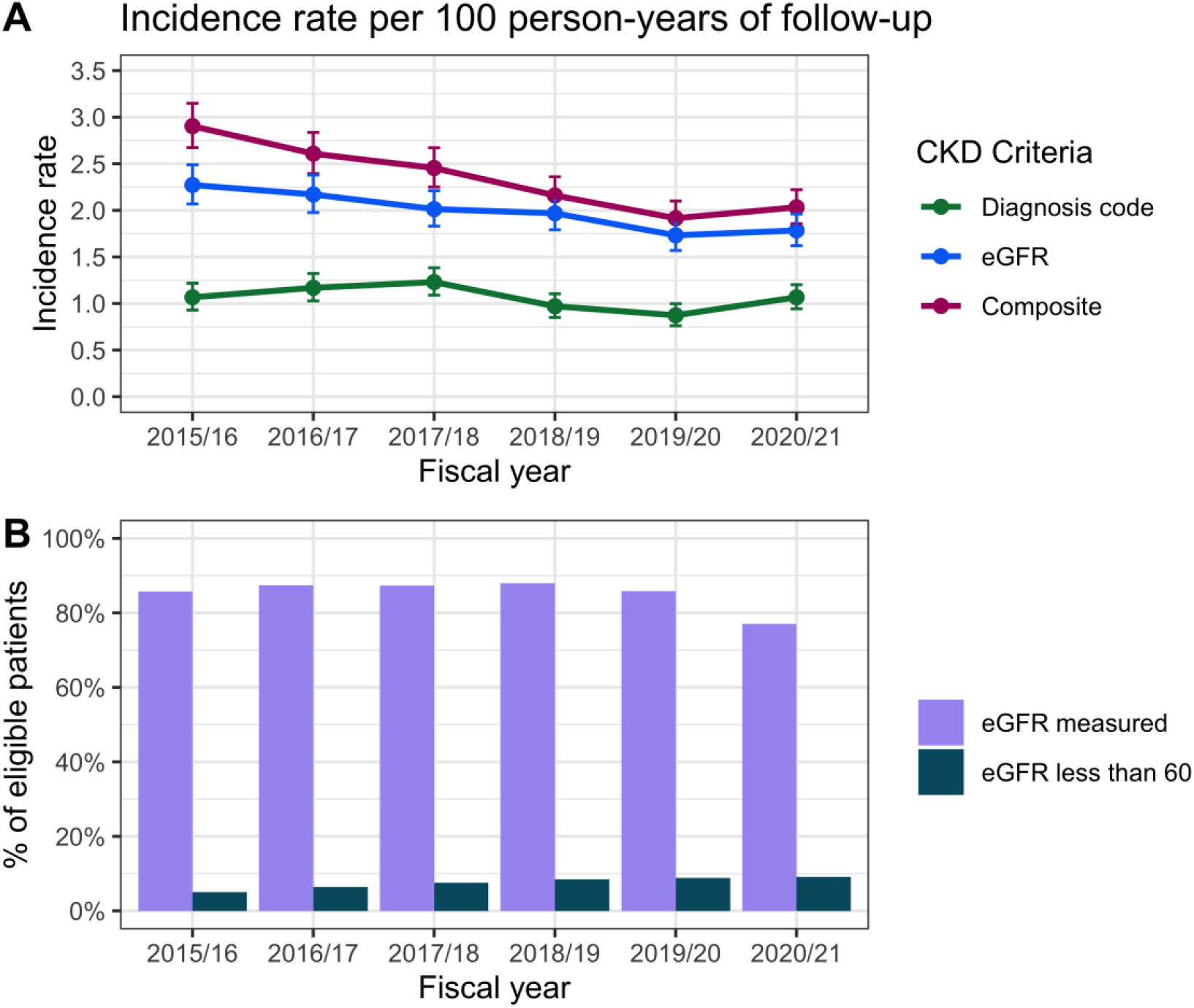
Amongst people with type 2 diabetes and no previous evidence of CKD stage G3-5, (A) The incidence rates of CKD diagnosis via a diagnosis code (Code), eGFR-based criteria (Lab) or at least one of these (Either), and their 95% confidence intervals. (B) Rates of eGFR measurement, by fiscal year

Few diagnosis code cases did not have evidence of eGFR-based criteria (N=411, 28.1%). However, a large proportion of eGFR-based cases did not have a corresponding diagnosis code (N=1,457, 54.6%). These findings were consistent over time (supplementary Table S**5**).

There was a statistically significant drop (p<0.001) in both the proportion of patients receiving at least one eGFR (serum creatinine) measurement in the 2020/21 fiscal year to 77.0% (95% CI: 76.5%, 77.5%), from 85.8% (95% CI: 85.4%, 86.2%) in 2019/20. Prior to 2019/20, the proportion of eligible patients with at least one valid eGFR or serum creatinine measurement remained high (Figure 2B). The proportion of patients with an eGFR less than 60 *ml/min/1.73m*^2^steadily increases over time from 5.1% (95% CI: 4.8%, 5.4%) in 2015/16 to 9.1% (95% CI: 8.7%, 9.4%) in 2020/21 (Figure 2B).

### 5.2. Diagnostic coding of patients meeting eGFR-based criteria

In total, 2,667 patients (8.3%) met the eGFR-based criteria for CKD during the study period. Of these, 54.6% patients did not have a corresponding diagnostic code, either before or after meeting the eGFR-based criteria (Table **2**).

**Table 2:** Rates of diagnostic coding, summarised by key patient characteristics. Unless specified otherwise, measures are calculated at the first qualifying eGFR measurement

|  | CKD Coded, N = 1,210 (45%)<sup>1</sup> | No CKD Code, N = 1,457 (55%)<sup>1</sup> | Overall, N = 2,667<sup>1</sup> | p-value<sup>2</sup> |
| --- | --- | --- | --- | --- |
| **Follow-up time after eGFR-based diagnosis (years)** | 3.20 (1.77, 4.70) | 2.38 (1.14, 3.92) | 2.75 (1.40, 4.36) | <0.001 |
| **Age (at index)** | 73.4 (66.8, 79.7) | 75.8 (69.8, 80.8) | 75.2 (68.1, 80.4) | <0.001 |
| **Gender, N (%)** |  |  |  | 0.3 |
| Female | 566 (47%) | 715 (49%) | 1,281 (48%) |  |
| Male | 644 (53%) | 742 (51%) | 1,386 (52%) |  |
| **Deprivation decile, N (%)** |  |  |  | >0.9 |
| 1-2 | 446 (37%) | 539 (37%) | 985 (37%) |  |
| 3-4 | 216 (18%) | 273 (19%) | 489 (18%) |  |
| 5-6 | 146 (12%) | 175 (12%) | 321 (12%) |  |
| 7-10 | 402 (33%) | 470 (32%) | 872 (33%) |  |
| **Duration of diabetes (years) at index** | 8.9 (4.6, 14.4) | 9.0 (4.3, 14.7) | 8.9 (4.4, 14.6) | 0.7 |
| **First qualifying eGFR (ml/min/1.73m<sup>2</sup>)** | 55 (49, 58) | 55 (50, 58) | 55 (50, 58) | 0.011 |
| **CKD Stage, N (%)** |  |  |  | 0.040 |
| G3A | 1,025 (85%) | 1,283 (88%) | 2,308 (87%) |  |
| G3B | 139 (11%) | 140 (9.6%) | 279 (10%) |  |
| G4 | 38 (3.1%) | 26 (1.8%) | 64 (2.4%) |  |
| G5 | 8 (0.7%) | 8 (0.5%) | 16 (0.6%) |  |
| **Time from index to qualifying eGFR (years)** | 0.56 (0.37, 0.87) | 0.59 (0.40, 0.90) | 0.57 (0.38, 0.89) | 0.035 |
| **Number of eGFRs between qualifying observations** | 2 (1, 3) | 2 (1, 3) | 2 (1, 3) | 0.4 |
| **Urine albumin-to-creatinine ratio (mg/mmol)** | 2.2 (1.1, 7.6) | 2.2 (1.0, 6.9) | 2.2 (1.0, 7.2) | 0.9 |
| **Urine albumin-to-creatinine ratio category, N (%)** |  |  |  | 0.067 |
| Not measured | 590 (49%) | 759 (52%) | 1,349 (51%) |  |
| A1 | 371 (31%) | 412 (28%) | 783 (29%) |  |
| A2 | 176 (15%) | 224 (15%) | 400 (15%) |  |
| A3 | 73 (6.0%) | 62 (4.3%) | 135 (5.1%) |  |
| **HbA1c (mmol/mol)** | 53 (46, 64) | 52 (46, 61) | 53 (46, 62) | 0.013 |
| **HbA1c measured in year prior to first qualifying eGFR** | 999 (83%) | 1,250 (86%) | 2,249 (84%) | 0.026 |
| **Prescribed antihypertensives, N (%)** | 1,006 (83%) | 1,204 (83%) | 2,210 (83%) | 0.8 |
| **Prescribed statins, N (%)** | 729 (60%) | 895 (61%) | 1,624 (61%) | 0.6 |
| **Prescribed insulin, N (%)** | 281 (23%) | 258 (18%) | 539 (20%) | <0.001 |
| **Prescribed SGLT2i, N (%)** | 39 (3.2%) | 40 (2.7%) | 79 (3.0%) | 0.5 |

<sup>1</sup>Median (IQR); n (%)
<sup>2</sup>Wilcoxon rank sum test; Pearson's Chi-squared test

Patients that had eGFR-based evidence of CKD were more likely to have a diagnosis code if they were younger, had an observed UACR (stage A1), or a higher G-stage of CKD at the first qualifying eGFR observation (Table **3**).

**Table 3:** Odds ratios (OR) and 95% confidence intervals from a logistic regression model to identify factors associated with diagnostic coding of eGFR-based cases

| Variable | Group | OR (95% CI) | p-value |
| --- | --- | --- | --- |
| **Age** |  | 0.973 (0.964, 0.982) | 0.00 |
| **Gender** | Female (ref) |  |  |
|  | Male | 1.067 (0.913, 1.247) | 0.41 |
| **Duration of diabetes (years)** |  | 1.000 (0.992, 1.009) | 0.91 |
| **Deprivation decile** | 1-2 (ref) |  |  |
|  | 3-4 | 1.094 (0.950, 1.260) | 0.21 |
|  | 5-6 | 1.022 (0.861, 1.214) | 0.80 |
|  | 7-10 | 0.931 (0.764, 1.134) | 0.48 |
| **eGFR Stage** | G3A (ref) |  |  |
|  | G3B | 1.219 (0.945, 1.572) | 0.13 |
|  | G4 | 1.798 (1.077, 3.048) | 0.03 |
|  | G5 | 1.043 (0.375, 2.898) | 0.93 |
| **Time between qualifying eGFRs (months)** |  | 0.848 (0.644, 1.115) | 0.24 |
| **Urine albumin to creatinine ratio (UACR)** | Not measured (ref) |  |  |
|  | A1 | 1.256 (1.048, 1.506) | 0.01 |
|  | A2 | 1.038 (0.826, 1.304) | 0.75 |
|  | A3 | 1.263 (0.876, 1.824) | 0.21 |
| **Prescribed antihypertensives** |  | 1.010 (0.821, 1.243) | 0.92 |
| **Prescribed statins** |  | 0.961 (0.819, 1.128) | 0.63 |
| **Prescribed insulin** |  | 1.217 (0.990, 1.496) | 0.06 |
| **Prescribed SGLT2i** |  | 0.903 (0.567, 1.435) | 0.67 |

### 5.3. Timeliness of diagnosis coding

The majority of patients (55.2%) with both eGFR-based CKD and a CKD diagnosis code received their CKD diagnosis code after their second qualifying eGFR measurement (Table **4**), and the median time from the second qualifying eGFR to entry of a diagnosis code was 9.79 months (IQR: 1.18, 24.34). 23.8% of patients with clinician-verified CKD received a diagnosis code within 30 days of meeting the eGFR-based criteria, 31.6% within 90 days, and 40.1% and 56.1% within 6 and 12 months respectively.

**Table 4:** Number of patients with eGFR-based CKD and a CKD diagnosis code, the proportion that occurred on or after their second qualifying eGFR measurement and the time between the second qualifying eGFR measurement and entry of a diagnostic code.

| Variable | Group | Number of Patients | Diagnosis Code After eGFR-based CKD, N (%) | Time from eGFR-based CKD to CKD Diagnosis Code (months), median [LQ, UQ] |
| --- | --- | --- | --- | --- |
| **Age group** |  |  |  |  |
|  | Under 40 | 3 | 1 (33.3%) | 0.7 [0.7, 0.7] |
|  | 40-49 | 19 | 11 (57.9%) | 2.9 [1.9, 9.6] |
|  | 50-59 | 111 | 56 (50.5%) | 8.9 [1.1, 16.3] |
|  | 60-69 | 298 | 156 (52.3%) | 10.9 [2.0, 28.2] |
|  | 70-79 | 496 | 293 (59.1%) | 9.9 [1.0, 24.8] |
|  | 80+ | 283 | 151 (53.4%) | 8.9 [0.9, 20.6] |
| **Gender** |  |  |  |  |
|  | Female | 566 | 319 (56.4%) | 10.6 [1.1, 24.1] |
|  | Male | 644 | 349 (54.2%) | 9.0 [1.2, 24.3] |
| **Duration of T2DM** |  |  |  |  |
|  | < 1 year | 77 | 48 (62.3%) | 11.8 [0.8, 26.2] |
|  | 1-2 years | 69 | 44 (63.8%) | 10.1 [2.0, 27.2] |
|  | 2-5 years | 177 | 94 (53.1%) | 7.6 [0.7, 26.1] |
|  | 5-10 years | 359 | 179 (49.9%) | 9.7 [1.1, 21.4] |
|  | 10+ years | 528 | 303 (57.4%) | 10.2 [1.5, 23.2] |
| **CKD Stage** |  |  |  |  |
|  | G3A | 1,025 | 569 (55.5%) | 9.9 [1.2, 25.3] |
|  | G3B | 139 | 78 (56.1%) | 10.2 [1.2, 20.4] |
|  | G4 | 38 | 17 (44.7%) | 7.3 [0.7, 16.6] |
|  | G5 | 8 | 4 (50.0%) | 7.2 [2.8, 21.5] |
| **Total** | Overall | 1,210 | 668 (55.2%) | 9.8 [1.2, 24.3] |

In total, 469 patients had at least one additional eGFR measurement between their CKD-qualifying eGFR less than 60 and entry of a CKD diagnosis code. At least one measurement of an eGFR less than 60ml/min/1.732 was observed in 4,351 patients. However, only 3,000 (68.9%) had a follow-up eGFR (or serum creatinine) within 6 months of their first abnormal result (supplementary table S3). 94 patients had an eGFR on the date of diagnosis coding, and these were on average 2.36 *ml/min/1.73m*^2^lower than their CKD-qualifying eGFR (supplementary figure S**5**, p = 0.019).

## 6. Discussion

### 6.1. Summary of findings

This analysis has provided concerning evidence of a lack of diagnosis coding of CKD in patients with type 2 diabetes in UK primary care, with more than half of patients meeting the eGFR-based criteria for CKD never receiving a diagnosis code. Of those patients that have a CKD diagnosis code after meeting the eGFR-based criteria, the entry of the diagnosis code occurs with a median delay of more than 9 months. Incidence was severely underestimated when using CKD diagnosis codes alone, which prevents reliable quantification of epidemiological estimates and the associated burden of CKD.

### 6.2. Relation to existing literature

Similarly to González-Pérez et al (20), we have presented crude estimates of CKD incidence per 100 person-years of follow-up by ascertainment criteria (eGFR-based CKD, diagnosis coded CKD or either). However, our combined estimates were considerably lower, likely due to several important differences in our criteria definitions; our study does not identify cases using UACR or albuminuria measurements, which accounted for 49% of González-Pérez et al’s included incidences. Furthermore, their work did not impose an upper limit on the time between qualifying eGFR measurements.

Our reported rates of diagnosis coding were lower than those presented by Molokhia et al(17), who observed that 57.5% of eGFR-based CKD cases in T2D patients in UK primary care had a diagnosis code. However, their data was limited to a single London borough extracted in 2013, their definition of eGFR-based CKD had no upper limit on the time between qualifying eGFRs, and they studied prevalence rather than incidence. Jain et al (16) conducted a similar study to establish whether patients with eGFR-based CKD were included on practice CKD registers, defined using QOF business rules and codelists. They observed that 77.7% of diabetic patients with eGFR-based CKD had a diagnosis code, which is higher than our reported coding rate. However, their study covered an earlier and shorter timeframe (April 2008 to March 2009) and they used a shorter length of time to identify sustained loss of function (two eGFRs < 60/*ml*/*min*/1.73*m*^2^ at least 7 days apart) that does not correspond to current guidelines. Further, they focused on prevalence rather than incidence.

### 6.3. Strengths and Limitations

To the best of our knowledge, this is the first study to quantify the rates of CKD diagnosis coding for new incidences of CKD in a type 2 diabetic population in UK primary care. We provide an analysis of recent data that covers the onset of the COVID-19 pandemic and therefore any associated impact on the management and identification of CKD. We hypothesise that the observed drop in eGFR measurement rates can be attributed to disruptions to the provision of routine care caused by the COVID-19 pandemic(21).

A limitation to this work is that our data only covers primary care. Patients may be referred to and managed within specialist secondary care centres upon exhibition of kidney function impairment. However, these referrals should be evidenced within the patient’s primary care record. Future work should explore whether and when such referrals take place, and establish how patients diagnosed with CKD in primary care are managed across the care pathway.

A further limitation is that our lab-based CKD definition used only eGFR. Albuminuria and/or an elevated UACR is often an earlier signal for kidney damage than a reduction in kidney filtration and is common in patients with type 2 diabetes. Therefore, further work should also capture patients that present with persistent albuminuria. However, it is well-known that adherence to UACR measurement guidelines is low, so incidence estimates based on UACR derived using routinely collected data are likely to be unreliable.

A final limitation is that the newest CKD-epi equation was used to calculate eGFR, without racial adjustment, which may provide a marginal overestimation of eGFR compared with what may have been calculated in practice. This may have resulted in comparatively fewer identified cases of eGFR-based CKD, however we do not anticipate that this would have a significant impact on the reported estimates since only the subset of patients close to the cut-off of eGFR < 60/*ml*/*min*/1.73*m*^2^would be affected.

### 6.4. Implications for clinical practice

A lack of, or delay in, identification and diagnosis coding of CKD could lead to improper management of the condition. CKD progression is strongly associated with poor clinical outcomes and has a significant economic burden. CKD awareness remains profoundly low, in part because CKD is usually silent until its late stages. Physician awareness of CKD is critical in the early identification of the condition as well as the early implementation of evidence-based therapies that can slow progression of kidney dysfunction, prevent metabolic complications, and reduce cardiovascular-related outcomes. Tools including automated CKD patient registry/diagnostic coding within electronic health records to identify and prioritise patients for early intensive management can facilitate the clinical inertia we see in CKD management. (22).

## Data Availability

Access to underlying patient data cannot be provided.

## 7. Conclusion

This study has provided evidence that CKD is poorly coded in primary care for people with type 2 diabetes, which could lead to improper care and delayed intervention. The reasons for poor coding require further investigation, and emphasis should be placed on examining historic test results for CKD to improve diagnosis coding.

## 9. Ethical approval

No ethical approval was required since only retrospective, anonymised data was used and therefore no direct patient contact or individual-level consent was necessary. The wider data collection and related analysis have been approved by the Health Research Authority (IRAS project ID: 288473).

## 11. Acknowledgements

We would like to acknowledge participating primary care centres across England that have contributed anonymised patient data that has been used in this work.

