## Supplemental materials for "Diagnosis coding of Chronic Kidney Disease in Type 2 Diabetes in UK primary care"

### Supplementary materials

#### Incidence, follow-up time and number of at-risk patients by definition and fiscal year

| | Composite CKD | | | | eGFR-based CKD | | | | | CKD Diagnosis Code | | | | |
|---|---|---|---|---|---|---|---|---|---|---|---|---|---|---|
| **Fiscal Year** | **N at-risk** | **N Inc.** | **Total FU** | **Inc. Rate** | **N at-risk** | **Total FU** | **N Inc.** | **Inc. Rate** | **p*** | **N at-risk** | **Total FU** | **N Inc.** | **Inc. Rate** | **p*** |
| 2015/16 | 21,856 | 585 | 20,144.7 | 2.90 (2.67, 3.15) | 21,856 | 20,201.0 | 459 | 2.27 (2.07, 2.49) | <0.001 | 21,856 | 20,335.0 | 217 | 1.07 (0.93, 1.22) | <0.001 |
| 2016/17 | 22,641 | 547 | 20,969.1 | 2.61 (2.39, 2.84) | 22,765 | 21,144.5 | 459 | 2.17 (1.98, 2.38) | <0.001 | 23,006 | 21,472.2 | 251 | 1.17 (1.03, 1.32) | <0.001 |
| 2017/18 | 23,618 | 534 | 21,753.7 | 2.45 (2.25, 2.67) | 23,821 | 21,997.6 | 443 | 2.01 (1.83, 2.21) | <0.001 | 24,265 | 22,515.4 | 277 | 1.23 (1.09, 1.38) | <0.001 |
| 2018/19 | 24,646 | 493 | 22,819.1 | 2.16 (1.97, 2.36) | 24,926 | 23,111.8 | 455 | 1.97 (1.79, 2.16) | 0.2 | 25,504 | 23,786.0 | 231 | 0.97 (0.85, 1.10) | <0.001 |
| 2019/20 | 25,500 | 454 | 23,700.0 | 1.92 (1.74, 2.10) | 25,809 | 24,014.7 | 416 | 1.73 (1.57, 1.91) | 0.1 | 26,546 | 24,831.7 | 217 | 0.87 (0.76, 1.00) | <0.001 |
| 2020/21 | 25,825 | 489 | 24,050.3 | 2.03 (1.86, 2.22) | 26,144 | 24,374.7 | 435 | 1.78 (1.62, 1.96) | 0.1 | 27,028 | 25,305.9 | 270 | 1.07 (0.94, 1.20) | <0.001 |
| **Overall** | **32,276** | **3,102** | **133,437.0** | **2.32 (2.24, 2.41)** | **32,276** | **134,844.4** | **2,667** | **1.98 (1.90, 2.05)** | **<0.001** | **32,276** | **138,246.2** | **1,463** | **1.06 (1.00, 1.11)** | **<0.001** |

***Table S1:*** *Raw incidence numbers (N Inc.), follow-up time in years (Total FU), number of at-risk patients (N at-risk) and incidence rates (Inc. Rate) per 100 person-years (and their 95% CIs) by definition and fiscal year. *P-value for the statistical differences between the incidence rates for eGFR-based CKD and CKD Diagnosis Code vs the composite CKD criteria (i.e. evidence of eGFR-based CKD or a CKD diagnosis code).*

### Occurrence and timing of follow-up measurements after the first observed eGFR < 60

| | N (%) / Median (IQR) |
|---|---|
| N patients with an eGFR < 60 | 4351 |
| Median (IQR) time until follow-up eGFR measurement (weeks) | 13.00 (3.43, 31.29) |
| N (%) patients followed up within 12 weeks | 2103 (48.3%) |
| N (%) patients followed up within 6 months | 3000 (68.9%) |

***Table S2:*** *Timing of follow-up after an observed eGFR < 60*

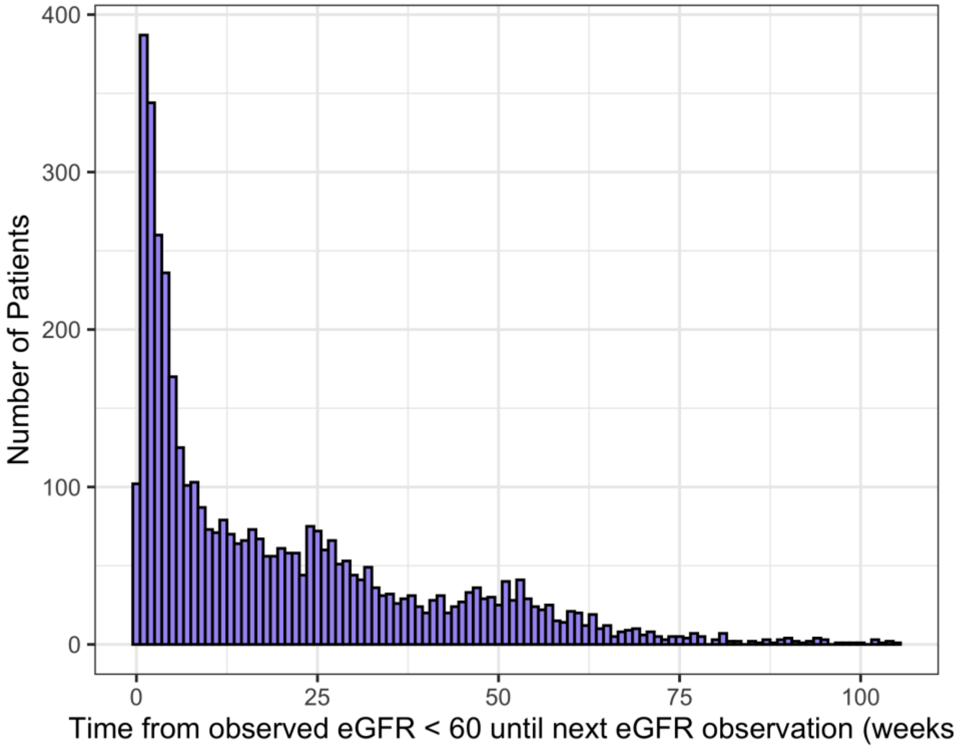


***Figure S1:*** *Histogram of time (in weeks) until the next eGFR observation for patients with an observed eGFR < 60. Limited to patients with at least 6 months of follow-up after observing an eGFR < 60, and with a maximum of 105 weeks until next observed eGFR.*

#### Histogram of time between qualifying eGFRs for eGFR-based CKD cases

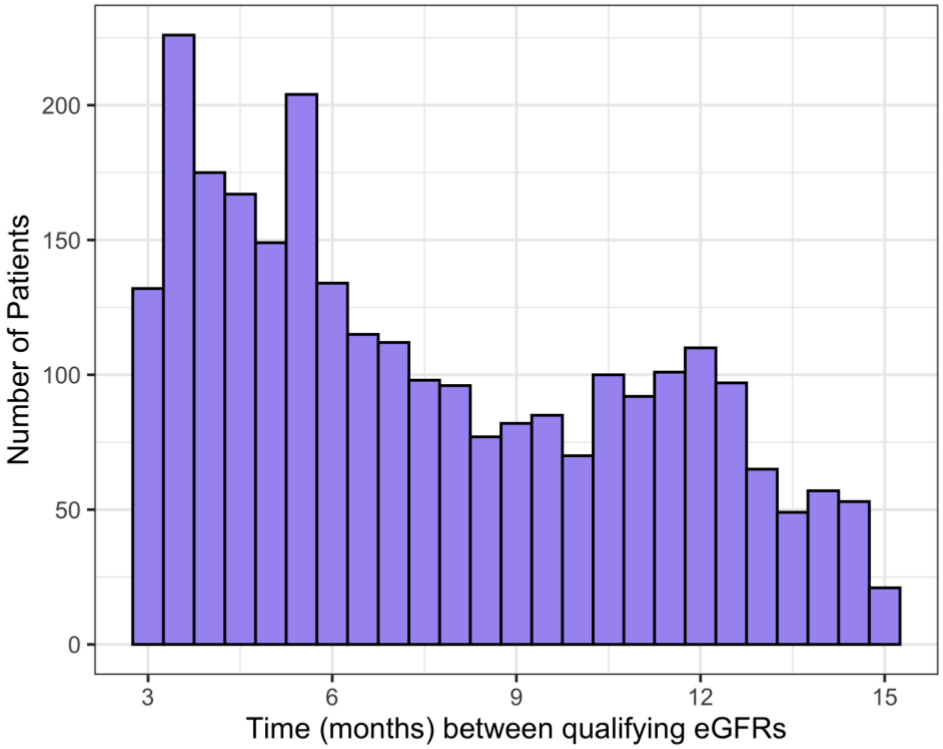


***Figure S2:*** *Distribution of time between qualifying eGFR measures for patients with eGFR-based CKD.*

#### Scatterplot of change in eGFR from CKD-qualifying measurement to entry of a diagnosis code

Figure **S3** shows the change in eGFR from a CKD-qualifying eGFR to entry of a diagnosis code with the corresponding time interval between the qualifying eGFR measurement and diagnosis coding. Only patients with an observed eGFR on the day of diagnosis coding are included (n = 94 patients).

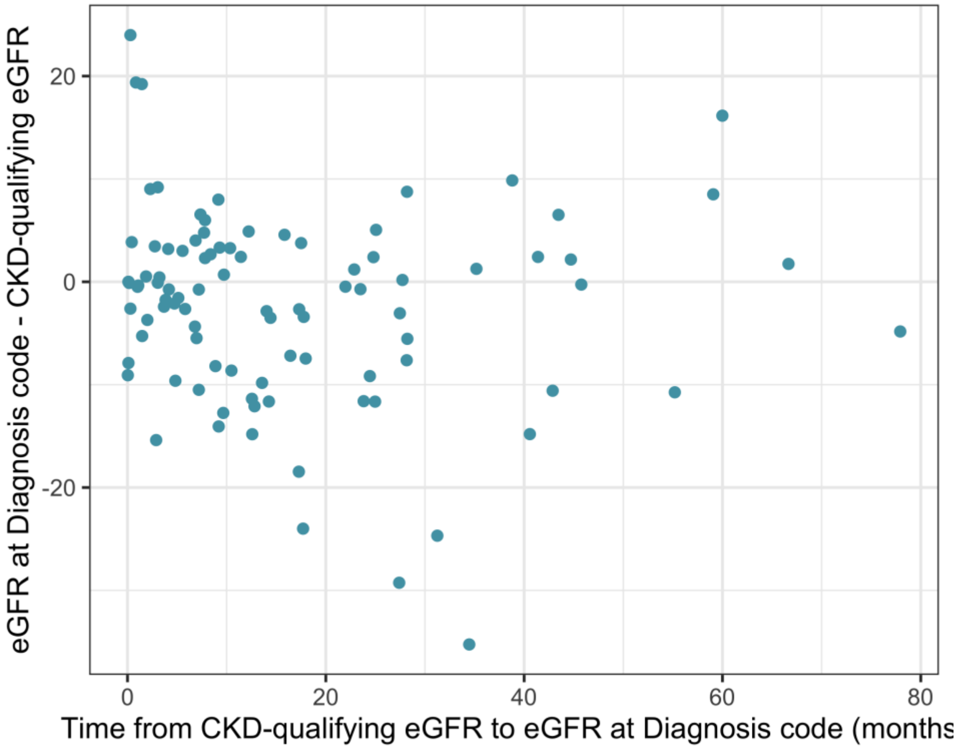


***Figure S3:*** *Change in eGFR from eGFR-based CKD to entry of a diagnosis code*

#### Code lists for Read V2, Read CTV3 and SNOMED terminologies

| Read V2 Code | Read Term | Stage |
|---|---|---|
| 1Z12. | Chronic kidney disease stage 3 | 3 |
| 1Z13. | Chronic kidney disease stage 4 | 4 |
| 1Z14. | Chronic kidney disease stage 5 | 5 |
| 1Z15. | Chronic kidney disease stage 3A | 3 |
| 1Z16. | Chronic kidney disease stage 3B | 3 |
| 1Z1a. | CKD G4A1 - chronic kidney disease with glomerular filtration rate category G4 and albuminuria category A1 | 4 |
| 1Z1b. | CKD G4A2 - chronic kidney disease with glomerular filtration rate category G4 and albuminuria category A2 | 4 |
| 1Z1B. | CKD stage 3 with proteinuria | 3 |
| 1Z1c. | CKD G4A3 - chronic kidney disease with glomerular filtration rate category G4 and albuminuria category A3 | 4 |
| 1Z1C. | CKD stage 3 without proteinuria | 3 |
| 1Z1D. | Chronic kidney disease stage 3A with proteinuria | 3 |
| 1Z1e. | CKD G5A2 - chronic kidney disease with glomerular filtration rate category G5 and albuminuria category A2 | 5 |
| 1Z1E. | Chronic kidney disease stage 3A without proteinuria | 3 |
| 1Z1f. | CKD G5A3 - chronic kidney disease with glomerular filtration rate category G5 and albuminuria category A3 | 5 |
| 1Z1F. | CKD stage 3B with proteinuria | 3 |
| 1Z1G. | CKD stage 3B without proteinuria | 3 |
| 1Z1H. | CKD stage 4 with proteinuria | 4 |
| 1Z1J. | CKD stage 4 without proteinuria | 4 |
| 1Z1K. | Chronic kidney disease stage 5 with proteinuria | 5 |
| 1Z1L. | CKD stage 5 without proteinuria | 5 |
| 1Z1T. | CKD G3aA1 - chronic kidney disease with glomerular filtration rate category G3a and albuminuria category A1 | 3 |
| 1Z1V. | CKD G3aA2 - chronic kidney disease with glomerular filtration rate category G3a and albuminuria category A2 | 3 |
| 1Z1W. | CKD G3aA3 - chronic kidney disease with glomerular filtration rate category G3a and albuminuria category A3 | 3 |

| Read V2 Code | Read Term | Stage |
|---|---|---|
| 1Z1X. | CKD G3bA1 - chronic kidney disease with glomerular filtration rate category G3b and albuminuria category A1 | 3 |
| 1Z1Y. | CKD G3bA2 - chronic kidney disease with glomerular filtration rate category G3b and albuminuria category A2 | 3 |
| 1Z1Z. | CKD G3bA3 - chronic kidney disease with glomerular filtration rate category G3b and albuminuria category A3 | 3 |
| K053. | Chronic kidney disease stage 3 | 3 |
| K054. | Chronic kidney disease stage 4 | 4 |
| K055. | Chronic kidney disease stage 5 | 5 |

***Table S3:*** *Read v2 diagnostic codes used to identify cases of Chronic Kidney Disease stages 3-5*

| Read CTV3 Code | Read Term | Stage |
|---|---|---|
| XacAb | CKD with GFR category G4 & albuminuria category A1 | 4 |
| XacAd | CKD with GFR category G4 & albuminuria category A2 | 4 |
| XacAe | CKD with GFR category G4 & albuminuria category A3 | 4 |
| XacAf | CKD with GFR category G5 & albuminuria category A1 | 5 |
| XacAh | CKD with GFR category G5 & albuminuria category A2 | 5 |
| XacAi | CKD with GFR category G5 & albuminuria category A3 | 5 |
| XacAM | CKD with GFR category G3a & albuminuria category A1 | 3 |
| XacAN | CKD with GFR category G3a & albuminuria category A2 | 3 |
| XacAO | CKD with GFR category G3a & albuminuria category A3 | 3 |
| XacAV | CKD with GFR category G3b & albuminuria category A1 | 3 |
| XacAW | CKD with GFR category G3b & albuminuria category A2 | 3 |
| XacAX | CKD with GFR category G3b & albuminuria category A3 | 3 |
| XaLHI | Chronic kidney disease stage 3 | 3 |
| XaLHJ | Chronic kidney disease stage 4 | 4 |
| XaLHK | Chronic kidney disease stage 5 | 5 |
| XaNbn | Chronic kidney disease stage 3A | 3 |
| XaNbo | Chronic kidney disease stage 3B | 3 |
| XaO3t | Chronic kidney disease stage 3 with proteinuria | 3 |
| XaO3u | Chronic kidney disease stage 3 without proteinuria | 3 |
| XaO3v | Chronic kidney disease stage 3A with proteinuria | 3 |
| XaO3w | Chronic kidney disease stage 3A without proteinuria | 3 |
| XaO3x | Chronic kidney disease stage 3B with proteinuria | 3 |
| XaO3y | Chronic kidney disease stage 3B without proteinuria | 3 |
| XaO3z | Chronic kidney disease stage 4 with proteinuria | 4 |
| XaO40 | Chronic kidney disease stage 4 without proteinuria | 4 |
| XaO41 | Chronic kidney disease stage 5 with proteinuria | 5 |
| XaO42 | Chronic kidney disease stage 5 without proteinuria | 5 |

***Table S4:*** *Read CTV3 diagnostic codes used to identify cases of Chronic Kidney Disease stages 3-5*

| SNOMED Concept ID | SNOMED Term | Stage |
|---|---|---|
| 433144002 | Chronic kidney disease stage 3 | 3 |
| 431857002 | Chronic kidney disease stage 4 | 4 |
| 433146000 | Chronic kidney disease stage 5 | 5 |
| 700378005 | Chronic kidney disease stage 3A | 3 |
| 324371000000106 | CKD stage 3B with proteinuria | 3 |
| 324311000000101 | Chronic kidney disease stage 3A with proteinuria | 3 |
| 324411000000105 | CKD stage 3B without proteinuria | 3 |
| 324341000000100 | Chronic kidney disease stage 3A without proteinuria | 3 |
| 324411000000105 | Chronic kidney disease stage 3B without proteinuria | 3 |
| 324341000000100 | CKD stage 3A without proteinuria | 3 |
| 700379002 | Chronic kidney disease stage 3B | 3 |
| 324471000000100 | CKD stage 4 without proteinuria | 4 |
| 324311000000101 | CKD stage 3A with proteinuria | 3 |
| 324281000000104 | CKD stage 3 without proteinuria | 3 |
| 950081000000107 | CKD G3bA2 - chronic kidney disease with glomerular filtration rate category G3b and albuminuria category A2 | 3 |
| 324281000000104 | Chronic kidney disease stage 3 without proteinuria | 3 |
| 324471000000100 | Chronic kidney disease stage 4 without proteinuria | 4 |
| 949881000000106 | CKD G3aA1 - chronic kidney disease with glomerular filtration rate category G3a and albuminuria category A1 | 3 |
| 324441000000106 | CKD stage 4 with proteinuria | 4 |
| 324251000000105 | CKD stage 3 with proteinuria | 3 |
| 433146000 | CKD stage 5 | 5 |
| 950211000000107 | CKD G4A2 - chronic kidney disease with glomerular filtration rate category G4 and albuminuria category A2 | 4 |
| 949901000000109 | CKD G3aA2 - chronic kidney disease with glomerular filtration rate category G3a and albuminuria category A2 | 3 |
| 324371000000106 | CKD (chronic kidney disease) stage 3B with proteinuria | 3 |
| 950061000000103 | CKD G3bA1 - chronic kidney disease with glomerular filtration rate category G3b and albuminuria category A1 | 3 |

| SNOMED Concept ID | SNOMED Term | Stage |
|---|---|---|
| 950181000000106 | CKD G4A1 - chronic kidney disease with glomerular filtration rate category G4 and albuminuria category A1 | 4 |
| 950101000000101 | CKD G3bA3 - chronic kidney disease with glomerular filtration rate category G3b and albuminuria category A3 | 3 |
| 949921000000100 | CKD G3aA3 - chronic kidney disease with glomerular filtration rate category G3a and albuminuria category A3 | 3 |
| 324371000000106 | Chronic kidney disease stage 3B with proteinuria | 3 |
| 324251000000105 | Chronic kidney disease stage 3 with proteinuria | 3 |
| 324441000000106 | Chronic kidney disease stage 4 with proteinuria | 4 |
| 324541000000105 | CKD stage 5 without proteinuria | 5 |
| 324501000000107 | Chronic kidney disease stage 5 with proteinuria | 5 |
| 950231000000104 | CKD G4A3 - chronic kidney disease with glomerular filtration rate category G4 and albuminuria category A3 | 4 |
| 714152005 | Chronic kidney disease stage 5 on dialysis | 5 |
| 950311000000102 | CKD G5A3 - chronic kidney disease with glomerular filtration rate category G5 and albuminuria category A3 | 5 |
| 950291000000103 | CKD G5A2 - chronic kidney disease with glomerular filtration rate category G5 and albuminuria category A2 | 5 |
| 324501000000107 | CKD stage 5 with proteinuria | 5 |
| 324541000000105 | Chronic kidney disease stage 5 without proteinuria | 5 |
| 950251000000106 | CKD G5A1 - chronic kidney disease with glomerular filtration rate category G5 and albuminuria category A1 | 5 |

***Table S5:*** *SNOMED diagnostic codes used to identify cases of Chronic Kidney Disease stages 3-5*
